# Deep Transfer Learning-based COVID-19 prediction using Chest X-rays

**DOI:** 10.1101/2020.05.12.20099937

**Authors:** Saurabh Kumar, Shweta Mishra, Sunil Kumar Singh

## Abstract

The novel coronavirus disease (COVID-19) is spreading very rapidly across the globe because of its highly contagious nature, and is declared as a pandemic by world health organization (WHO). Scientists are endeavoring to ascertain the drugs for its efficacious treatment. Because, till now, no full-proof drug is available to cure this deadly disease. Therefore, identifying COVID-19 positive people and to quarantine them, can be an effective solution to control its spread. Many machine learning and deep learning techniques are being used quite effectively to classify positive and negative cases. In this work, a deep transfer learning-based model is proposed to classify the COVID-19 cases using chest X-rays or CT scan images of infected persons. The proposed model is based on the ensembling of DenseNet121 and SqueezeNet1.0, which is named as DeQueezeNet. The model can extract the importance of various influential features from the X-ray images, which are effectively used to classify the COVID-19 cases. The performance study of the proposed model depicts its effectiveness in terms of accuracy and precision. A comparative study has also been done with the recently published works and it is observed the performance of the proposed model is significantly better.

## 1. Introduction

COVID-19 stands for Coronavirus Disease-19, is emerged as an epidemic, which has attracted worldwide attention since December-2019. Initially, it was referred to as the novel coronavirus 2019 (2019-nCoV) but later on, was renamed as COVID-19 officially by the World Health Organization (WHO) on February 11, 2020 [1]. Coronavirus disease (COVID-19) is an infectious disease, caused by a new virus. This disease makes humans suffer from respiratory illness with symptoms like cough, high fever, and in severe cases, difficulty in breathing.

If we look at the history of coronaviruses, these are the viruses that cause disease in mammals and birds. In humans, coronaviruses cause respiratory tract infections (RTI) [2] that can vary from mild to lethiferous. The first virus of the family Coronaviridae was SARS-CoV, which stands for severe acute respiratory syndrome coronavirus. Its outburst started in Guangdong, China, and spread to many other countries in Southeast Asia. The last case of SARS-CoV was reported in September 2003 and it infected 8000 persons causing 774 deaths with a case mortality rate of 9.6%[1].

After nine years around in mid-2012, another deadly virus appeared in the Middle East from this family, which was known as Middle East respiratory syndrome coronavirus (MERS-CoV)[1, 3]. When we compare the case mortality rate of MERS-CoV with SARS-CoV, it is quite high and is around 35%.

COVID-19 is again a novel virus from the Coronaviridae family, which emerged from the City of Wuhan China [4], has spread to several other countries. It is a zoonotic origin virus i.e. transmitted from animals to humans; now it is transmitting between humans to humans.

Novel coronavirus or COVID-19 spread like a wildfire, wilder than the SARS in 2003. Figure 1 shows the worldwide infected cases at an interval of 10 days. The vertical axis represents the numbers of confirmed, recovered, and death cases. As shown in the figure, on the date **02.05.2020** number of confirmed, recovered and death case are **3,427,343**, **1,093,112** and **243,808** respectively.

**Figure 1.**
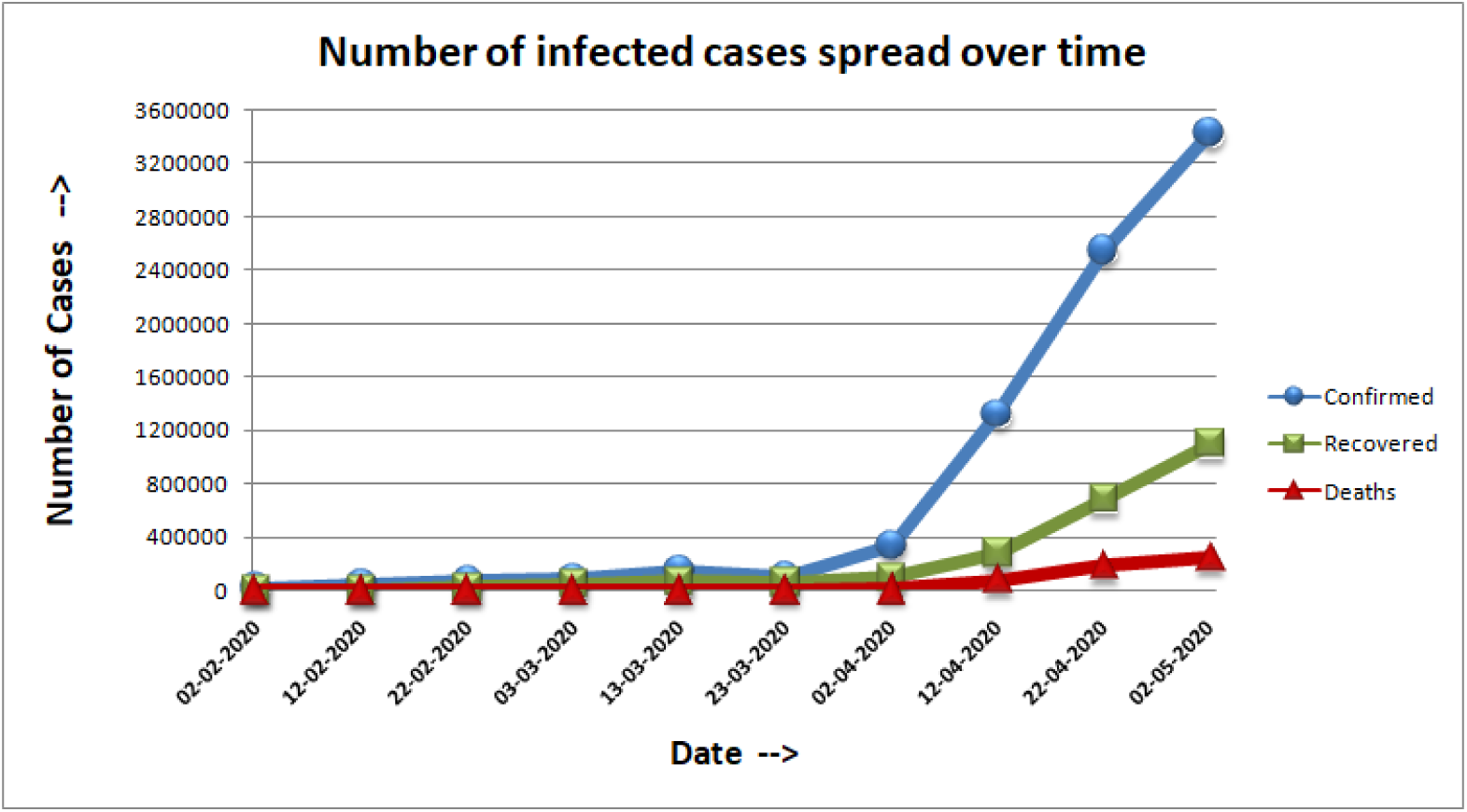
Spread rate of Worldwide infected cases

In the beginning, it increases slowly, when it is in the first phase of transmission, but once it reaches into third/community transmission phase (community transmission phase is the one in which source of infection cannot be traced) then the rate of increase in the number of cases is very high. For example, the number of confirmed cases can be observed on April 2^nd^,2020, and April 12^th^, 2020 which is around 1.01M and 1.83M respectively showing a steep increase of 800K+ cases in small tenure of 10 days. As per its widespread and worldwide impact, WHO declared COVID-19 as a pandemic on March 11^th^, 2020.

At the time of writing (May 7^th^, 2020), the worldwide number of confirmed, recovered, and fatal coronavirus cases are 3.7M+, 1.2M+, and 250K+ respectively and the mortality rate is more than 6.75%. If we look at the highly affected countries, leading the charts are United States of America (from the North-American region), Spain, Italy, Germany (from European region), China, Iran (from Eastern-Mediterranean region) and many other countries including India (from South-East Asia region). As per the WHO report, around the whole world is suffering from this damning disease, and its global risk assessment level is very high.

Unfortunately, until now, clinical features for identifying various stages of COVID-19 pneumonia [5] is still not clear, due to that, infected persons are often being identified after reaching into severe stages. Scientists are endeavoring to discover the drugs for their effective treatment. For the time being, Chloroquine and Hydroxychloroquine have been found effective for controlling the incisiveness of pneumonia, improving lung imaging findings, and promoting a virus-negative conversion [6, 7]. Still, it is not a satisfactory solution because the rapid increase in the number of cases can not be checked effectively and that leading to infect many others. Therefore, identifying the cases in early-stage and quarantine the infected person is going to be a relevant solution.

There are many machine learning and deep transfer learning-based models being applied to X-Ray images to detect COVID-19 cases effectively. But speedy and accurate detection is still a challenging task. For this, we have proposed a deep transfer based learning model; which is an ensemble of DenseNet121 and SqueezeNet1.0 to detect the COVID-19 cases effectively.

The remaining part of the paper is organized as follows. Section 2 highlights the related work which covers a few recently published models in the area. Section 3 and 4 states the problem statement and data acquisition, pre-processing respectively. Section 5 highlights the transfer learning, ensemble learning, fundamental architecture of DenseNet121, and SqueezeNet. It also defines the proposed model and its flow. The performance analysis of the proposed model is covered in section 6. Section 7 concludes the model and its importance.

## 2. Related work

A few similar works have been done in which many deep learning techniques have been applied. Applying these techniques over medical images such as X-Rays, CT Scans, MRI has been in practice since long and success rate is quite significant [8, 9]. The implementations of deep learning techniques have a wide range over multiple tasks such as disease diagnosis [10], survival rate prediction [11], tumor segmentation [12], and many other applications. DenseNet is also being used frequently in the field of medical imaging. Mentioning some noteworthy works, it is used for cardiac segmentation and diagnosis [13], a fully connected framework is used for semantic segmentation [14], a transfer learning-based framework is used on fundus medical images data [15], outcomes have been quite encouraging in all these cases.

In the proposed model, we have used X-Ray images for prediction, which also have a wide range of applications in deep learning, especially in case of pneumonia prediction, the symptoms of which are very similar to that of COVID-19. A few highly accurate and clinically approved uses of X-Rays in deep learning are, radiologist-level pneumonia detection, where architecture was constructed with name ChexNet by Rajpurkar et. al [16] and tuberculosis detection using deep convolutional neural network architecture called TX-CNN used in [17].

A few works have been done to diagnose the COVID-19 using machine learning and deep learning frameworks, to note some, Narin et al. [18] propose to use a deep convolutional network-based model called ResNet50 and compare its performance concerning two other models namely InceptionV3 and InceptionResNetV2 for detecting coronavirus infected patients using X-Ray images. Results establish that, among these three, ResNet50 is the best performing model in terms of accuracy. But the reliability of this model is questionable, as it is tested on a quite small data set and its performance needs to be observed on a larger dataset also.

Another noteworthy study has been performed by Sethy and Behra [18], where they have stacked a support vector machine (SVM) in front of ResNet50 to detect corona virus-infected persons using X-ray images.

A deep learning-based model to detect the disease from Chest CT scans using weak label [19] and an Inception Net inspired model also using CT scans to predict the disease [20], were also some recently published works on the same topic. Gaining motivation from all the aforementioned works, the proposed model aims to achieve higher efficiency standards on a larger dataset so that it can be used for clinical purposes.

## 3. Problem Statement

Currently, many countries of the world, including India are suffering from COVID-19. A few countries like the USA, Germany, Italy, and several others are facing its spread in the community transfer phase, which indicates that one infected person can infect more than 100 people to whom he contacts. So, the solution to the problem is to identify the infected persons and put them in quarantine to stop the further spread. Existing diagnosis procedures to identify the infected person, are time-consuming which is affecting the rate of diagnosis when dealing with a large number of cases.

Therefore, for overcoming this issue, we have proposed a model, which can efficiently classify the COVID-19 positive and negative cases well advance in time.

So, provided the problem, the task is to classify people as COVID-19 positive or COVID-19 negative, by using a suitable automated machine learning-based model. The model takes X-ray images as an input parameter which are showing initial symptoms of the disease.

## 4. Data

This section of the model states the data acquisition and discusses the pre-processing applied to the data.

### 4.1 Background and Data Acquisition

The diagnosis of COVID-19 is confirmed by polymerase chain reaction (PCR), which is a complex procedure comprising of heavy machinery and transfer requirement of test samples, consuming a lot of time. Also, various studies suggest that patients suffering from pneumonia can be diagnosed by observing the abnormalities in their respective chest X-rays or CT scans [21]. As we do in pneumonia-like diseases, the same procedure can be followed with the patients showing symptoms of COVID-19[22], which can save a lot of valuable time, as conducting X-rays is quite a feasible and quick procedure, we can conduct a large number of tests in a short time.

So, for our study, we have used the chest X-Ray images of patients obtained from COVID-19 image data collection [23], an open-source dataset. This dataset, at the time of the acquisition, was significantly small and highly skewed towards COVID-19 positive samples. So, to increase our sample size and make it skewed towards negative samples, transforming into a more realistic scenario, we have included the X-Ray images of either normal or pneumonia infected people from another open-source dataset [24] at Kaggle for the studies related to bacterial and viral pneumonia prediction.

### 4.2 Pre-Processing

The data collected from the aforementioned two sources is pre-processed to convert it into a usable form, in which first, we have scaled all the images of data set to a uniform size of 512 × 512 and then distributed the data into three parts, training, validation and test sets. The data set contains 401 images out of which, 262 images are COVID-19 negative and the remaining 139 are for positive cases. Firstly a set of 73 images was selected randomly for testing purposes. Out of these 73 selected images, 21 were COVID-19 positive instances and the remaining 52 were COVID-19 negative instances. The remaining images were randomly shuffled and then a 3:1 training-validation split was performed, producing 246 images for training and 82 for validation purposes.

## 5. The Model

The model section comprises an introduction to transfer learning, ensemble learning, DenseNet, SqeezeNet and it also describes the definition and detailed architecture of the proposed model and in the end, states the training pattern.

### 5.1 Transfer Learning

In modern machine-dependent automated prediction and classification tasks, deep learning is considered to be the most sought-after concept, thanks to the remarkable performance obtained by it. But it suffers from the problem of data-dependence, as, it’s models require a tremendous amount of data to train them, due to their complex architecture. The new find cases like COVID-19, have an insufficient amount of training data, making it difficult to use deep learning architectures for tasks related to them. Therefore, to deal with the problem of insufficient training data, an existing machine learning tool has been used, which helps us to transfer pre-existing knowledge from other tasks or the source domain to achieve the target for our concerning task or the target domain, this whole procedure is known as transfer learning [25].

In case of deep learning, given a very deep neural network for one generalized task of image classification, the initial layers and their weights are task-independent, only the final layers decide which kind of image classification task it would be, so we can easily import the weights of the initial layers of a pre-trained model and then train the final ones with whatever data we have, to make the model task-specific. This whole process is broadly termed as deep transfer learning.

### 5.2 Ensemble Learning

Ensemble learning, in the context of deep neural networks, is training different neural networks with the same input data configuration and measuring the average value of the predictions made by each network to have our final prediction.

There are several methods to take the average, such as mean and mode. For our model, we have used mean predictions.

Given *N* neural networks, each making a prediction *p_i_*, the final prediction P made by a mean-based ensemble method is defined as mentioned in equation 1.

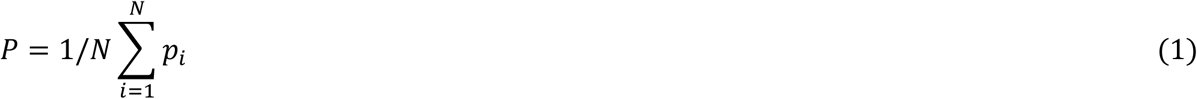

### 5.3 Model Selections to Ensemble

Using the same training, validation, and test data, we trained 5 deep transfer learning models namely AlexNet, DenseNet121, ResNet50, VGG16, and SqueezeNet1.0. The best performing models out of these 5 are DenseNet121 and SqueezeNet1.0. As indicated in figure 2, DenseNet121 and SqueezeNet1.0 return the highest precision, recall, and accuracy values out of all five models.

**Figure 2.**
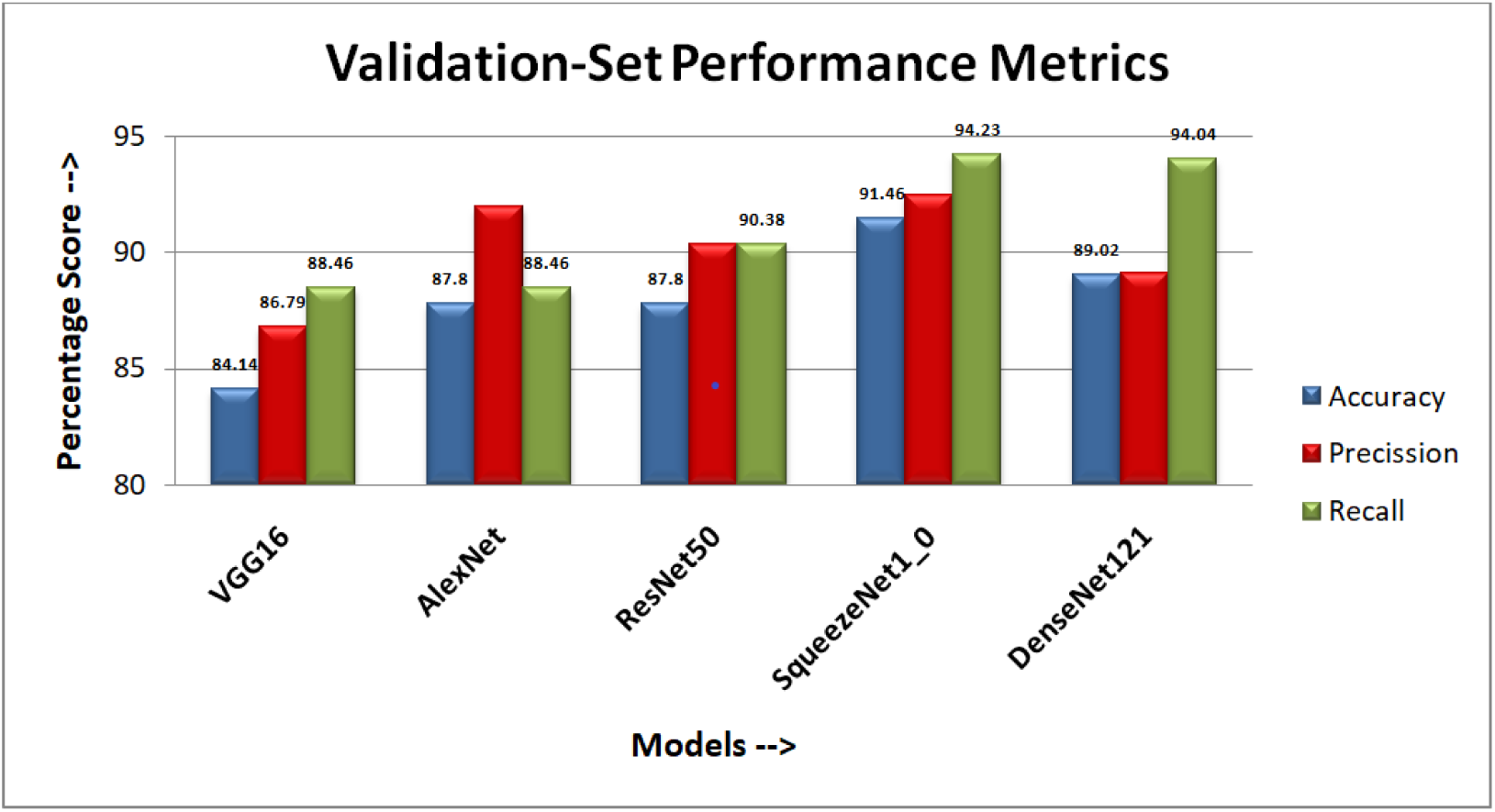
Validation-set performance metrics for model selection

Also, an uncanny relation was there in between these 2 models on results got on test-set, that wrong predictions made by the classifiers were mostly on different samples.

Hence, it is appropriate to take the ensemble of these two (DenseNet121 and SqueezeNet1.0) models, to compensate for the errors of prediction of each other.

### 5.4 DenseNet

For image classification tasks, Convolutional Neural Networks (CNNs) are the state-of-the-art neural architectures used all along with the world. With time, in an attempt to further improve the accuracy, researchers have made many alterations with its architecture. As the complexities of tasks have increased, therefore it is required to use deeper network architectures to process it effectively. Now, as the hardware availability has improved significantly, true deep CNN architectures are being proposed and trained. The barrier to the number of layers is being broken with time, be it 16 or 19 layers of VGG [26] or more than 100 layers in Highway Networks [27]. Also, a new trend is there to pass the residues of a layer to layers way ahead of them as in ResNet [28], i.e. a layer-1 will not only forward its feature map to layer-2 but can also forward it to layer-3, layer-4 or any forthcoming layer. Multiple architectures have more than 1000 layers in them.

To further improve the information flow between layers, it is proposed to connect each layer to all its subsequent layers i.e. for a given layer *l* the input *x_l_* is defined as in equation 1.

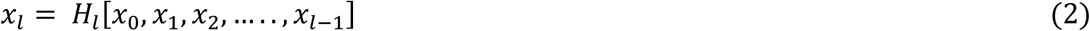

Where, *H_l_*[*x*_0_*,x*_1_*,x*_2_*,…..,x_l_*_−1_] is a combined feature map of all previous layers [0,1,2, *….,l* − 1].

This process enabled very dense connectivity between layers and the resulting network is termed as Dense Convolutional Network (DenseNet) [29].

Many DenseNet models are in use, here, DenseNet121 is used as one of the models for ensembling into the proposed model, where 121 is the total number of layers present in the architecture. The task flow in the DenseNet121 model can be visualized as shown in figure 3.

**Figure 3.**
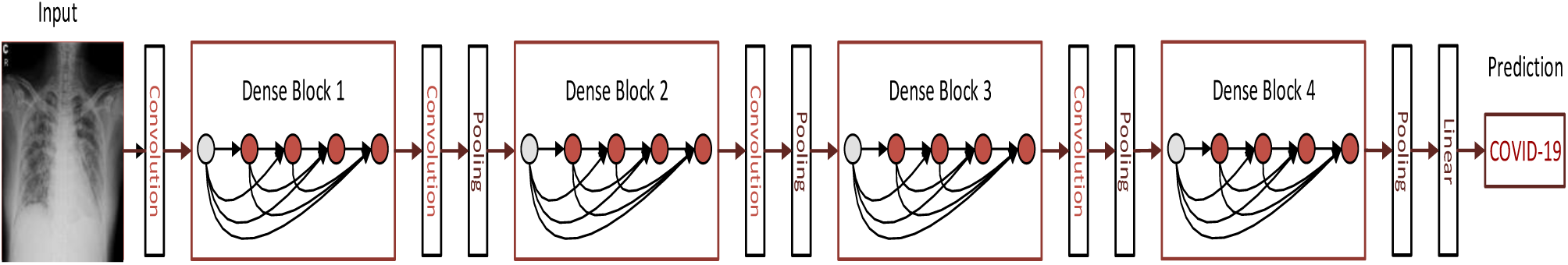
An Architecture of the DenseNet121 model

It is a deep DenseNet with four dense blocks, in which the layer between two adjacent blocks is referred to as transition layers. The transition layers are intended to change the feature-map sizes via convolution and pooling.

The architecture takes the image as input and after detection shows the output in the form of COVID-19 positive or negative.

### 5.5 SqueezeNet

This model also was a breakthrough in the field of true deep convolutional neural networks. The main goal achieved by this model was to gain AlexNet level accuracy while minimizing the number of parameters as compared to AlexNet to a great extent.

The model, in an uncompressed form, is consists of 50x fewer parameters than AlexNet, and do also maintains the same accuracy level as AlexNet, over ImageNet data.

In order to achieve the aforementioned goals, this model introduced a novel concept of fire module. The model uses 9 fire modules in total, each having two blocks, squeeze block, having 3 1×1 convolution filters, and an expand block having 3 1×1 as well as 4 3×3 convolution filters [30].

The first version of this model, named SqueezeNet1.0 has been used in this study.

### 5.6 The Proposed Model

The proposed model is an ensemble of DenseNet121 and SqueezNet1.0, in an attempt to increase the efficiency of Covid-19 prediction.

For this model, we separately train DenseNet121 and SqueezeNet1.0, and then on any given chest X-ray images from the data sample, take the mean of the prediction probabilities obtained from these two DenseNet121 and SqueezeNet models to get the final prediction. For any given chest X-Ray image I, the function of DenseNet121 and SqueezeNet1.0 can be expressed as follows.

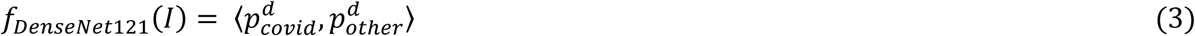

And,

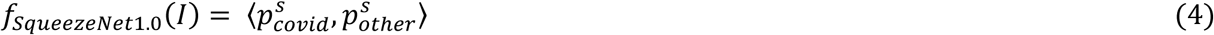

Equation 3 is the function for DenseNet121, with output 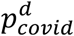 as the probability of *I* being a COVID-19 positive sample and 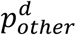 as the probability of *I* being a COVID-19 negative sample.

And equation 4 is the function of SqueezeNet1.0, with output 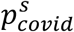 as the probability of *I* being a COVID-19 positive sample and 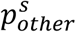 as the probability of *I* being a COVID-19 negative sample.

Now, for the proposed model i.e. DeQueezeNet, the probability of X-ray image *I* being COVID-19 positive is shown by equation 5.

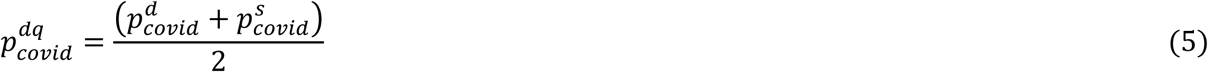

Where equation 6 indicates the probability of image I being COVID-19 negative.

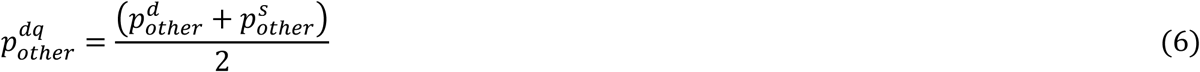

Hence the final predicted class of image *I* by DeQueezeNet (The proposed model), denoted by σ, returning either 0 (denoting COVID-19 positive class) or 1 (denoting COVID-19 negative class), can be written as mentioned in equation 7.

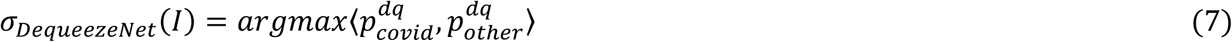

### 5.7 Flow chart of DeQueezeNet

Figure 4 shows the flow chart of the proposed model i.e. DeQueezeNet, which is an ensemble version of Squuezenet1.0 and DenseNet121.

**Figure 4.**
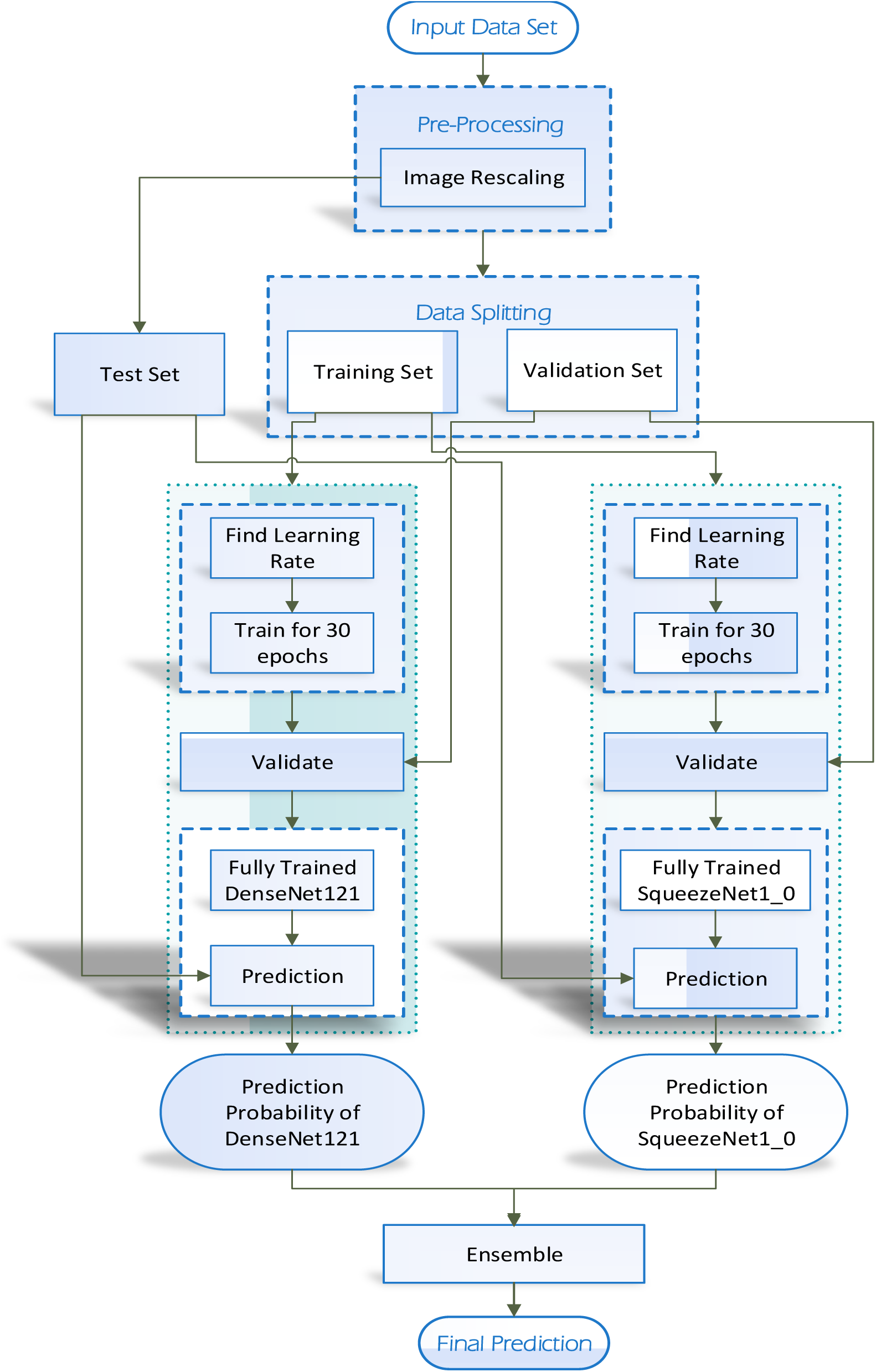
Flowchart of the proposed model

As shown in figure 4, we take the X-ray image dataset as input and resize all images as a part of pre-processing into 512 × 512 and reserve a fraction of the dataset [73 images] for testing purposes.

The rest part of the pre-processed data set is split into training and validation set, in which 75% of the data is for the training and the remaining 25% for the validation set.

Using the training dataset, we train DenseNet121 and SqueezeNet1.0 separately, followed by validation of the trained models.

Fully trained and validated models take the test set as input and predict the outcome in terms of probability. And these predicted probabilities are ensembled for further processing.

In the final step, the probability obtained after the ensemble is used to predict the COVID-19 positive and negative cases.

### 5.7 Model Training

The models used for the ensemble in the proposed are trained as follows.

Although there are multiple variants of DenseNet and SqueezeNet we have utilized DenseNet121 (121 is the number of layers present) and SqueezeNet1.0 (the first version of SqueezeNet) for our study.

Due to limited availability of data, transfer learning method has been used, so we imported a pre-trained model of DenseNet121 over, ImageNet, which is an image database consisting of millions of images organized according to the WordNet hierarchy where each node of the hierarchy is represented by a collection of images, averaging 500 images per node [31]. The model was imported from fastai library [32] and then our training and validation data were fed onto it.

For training both the models (DenseNet121 and SqueezeNet1.0), we have used the LR-finder module of fastai [30] to find the optimal learning rate suited for the training data. The LR-finder works by selecting a sequence of learning rates and training the models for 1 epoch each, using each learning rate. In the case of the high learning rate, loss of information starts to explode or increases exponentially. Therefore, admissible maximum learning rates are chosen for training the models.

In figure 5 and figure 6, the learning rate versus loss plot after running the LR-finder module is shown, with figure 5 denoting plot for DenseNet121 and figure 6 denoting plot for SqueezeNet1.0. From the observation, it is derived that the maximum permissible learning rate for the model DenseNet121 is 1*e*– 04 and for SqueezeNet1.0 is 2*e*–04 because, beyond these levels, the loss starts to increase exponentially. Both the models are trained for 30 epochs, using there aforementioned maximum learning rates utilizing fit-one-cycle policy [31]. In fit-one-cycle policy, the learning rate is adjusted slightly after completion of each epoch.

**Figure 5.**
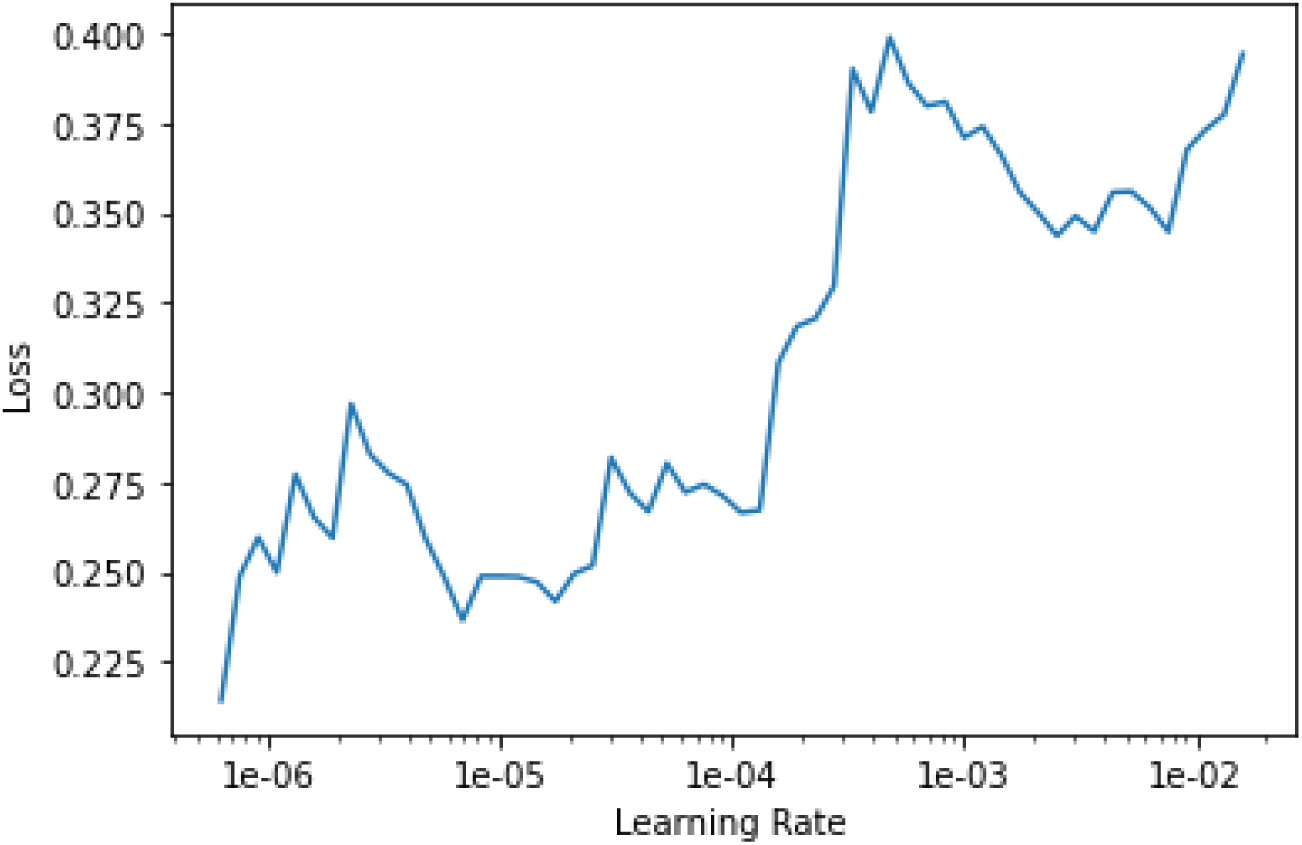
The outcome of LR-finder module on DenseNet121

**Figure 6.**
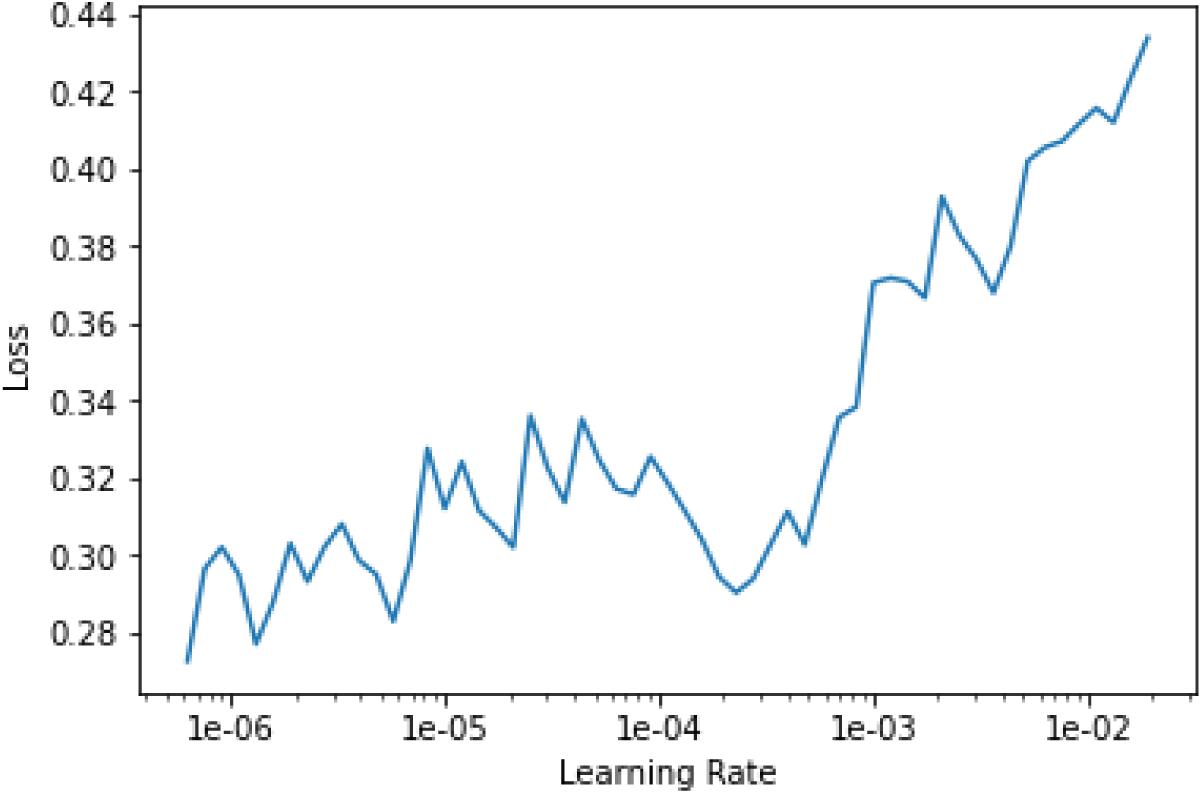
The outcome of LR-finder module on SqueezeNet 1.0

## 6. Performance Analysis and Comparative Study

For studying the performance of the proposed model, programming is done on Python-3 and the model is trained and tested on Google Colaboratory, a cloud-based programming platform.

Performance analysis of the proposed model is observed by plotting the confusion matrix for validation and test samples and the comparative study indicates the importance of the model.

### 6.1 Performance Metrics

A few important metrics used to compare the performance of the proposed model to other models are as follows.

#### 6.1.1 Losses on training and validation

The loss is basically calculated by computing the difference between the actual and predicted values of the class probabilities. While comparing the models based on their losses, we have to look for three factors; first, the loss should be low and its difference should be as low as possible, second, if the training loss is much lesser than the validation loss, then the corresponding model is overfitting, and the third, if the validation loss is lesser than training loss, then the corresponding model is underfitting. The loss function used in our model is Softmax, quite a popular function for calculating the loss in binary classification.

#### 6.1.2 Accuracy

Accuracy [33] is calculated as the percentage of samples correctly classified concerning the total number of samples as indicated in equation 8. In which, *S* is the total sample size, 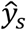 denotes the predicted label and *y_s_* denotes the ground truth label.

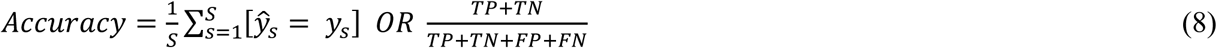

Where TP: true positive, TN: true negative, FP: false positive and FN is a false negative.

A higher accuracy value approves the effectiveness of the model.

#### 6.1.3 Precision

In machine learning, precision [33] shown in equation 9, is defined as the percentage of a specific class of samples correctly guessed by a model to the total number of samples of that very class. In the proposed model, the class is of COVID-19 positive people, which is considered for it.

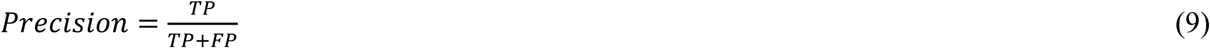

#### 6.1.4 Recall

Recall is defined as the percentage of actual positive samples predicted correctly. In the proposed model, it is calculated in the form of a percentage of COVID-19 positive samples correct predictions. It can be calculated with the help of equation 10.

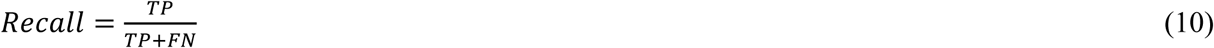

#### 6.1.5 Area under ROC curve (AUROC)

Receiver Operating Characteristics (ROC) curve [33] is a graphical representation of the true positive rate (TPR) plotted against the false positive rate (FPR). TPR and FPR can be calculated as given in equations 11 and 12. The higher TPR and lower FPR incur the high percentage area inside the plot, indicates, how much better a model is interpreted to be.

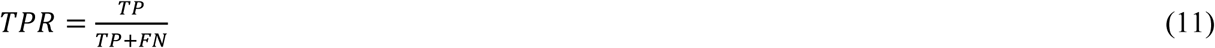

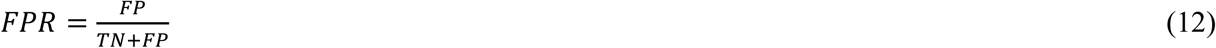

### 6.2 Confusion Matrix

The confusion matrix is shown to analyze the performance of the proposed model DequeezeNet. On the test set, Figure 7 shows the outcomes obtained by the DeQueezeNet model.

**Figure 7.**
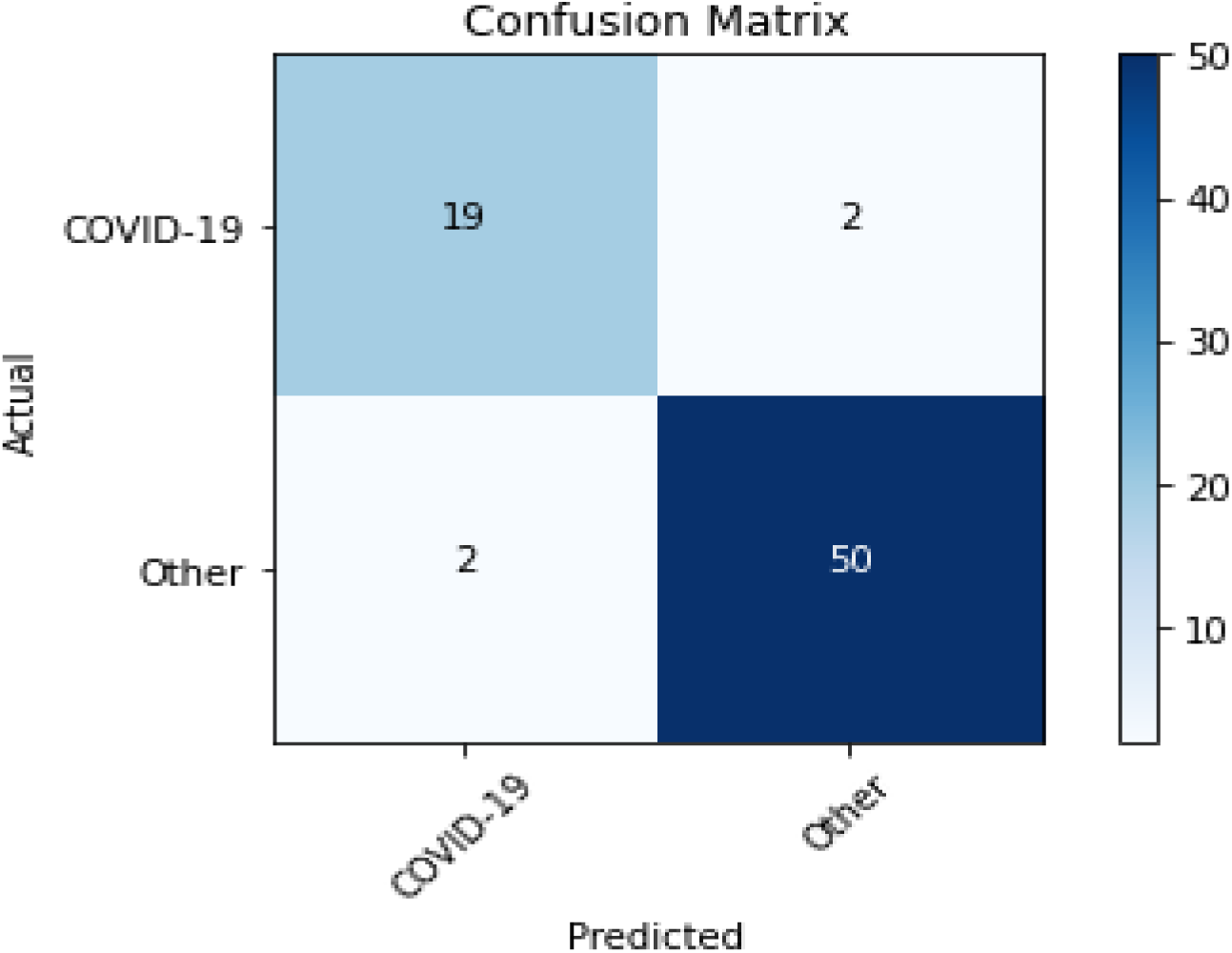
Confusion Matrix of proposed Model DeQueezeNet

Observation from the figure 7 shows that out of 21 positive samples, 19 were classified correctly, and out of 52 negative samples, 50 were classified correctly, making it 69 out of 73 correct predictions, with an accuracy of 94.52 %, precision of 90.48% and recall of 96.15%. All of which are pretty decent scores, emphasizing the usability of the model in practical and clinical cases.

### 6.3 Comparative Analysis

To study the comparative performance of the proposed model, we chose 3 different models for comparison, DenseNet121, SqueezeNet1.0 (both models have been used to build the proposed model DeQueezeNet), and ResNet50 is given by Narin et al. [34] for detecting the coronavirus disease using X-ray images.

These models are applied to predict the COVID-19 positive and negative cases, figure 8 shows the accuracy over the varying number of epochs from 1 to 30.

**Figure 8.**
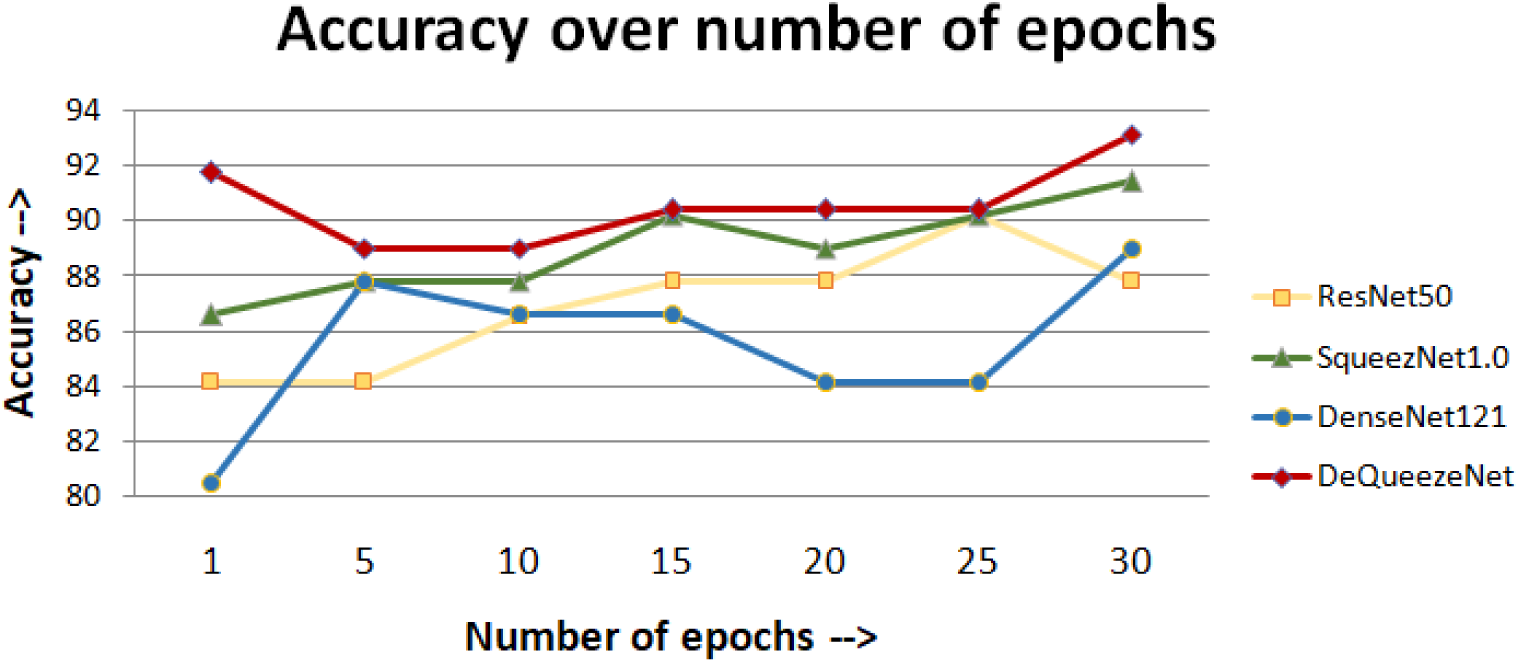
Comparative Accuracy over the number of epochs

Observation from figure 8 shows that the proposed model DeQueezeNet is performing better than all the other models for every epoch of training. The proposed model’s performance is better even when it is trained for just one epoch.

The second set of comparative study is done to observe the Percentage Score of the proposed model. All the models are trained up to 30 epochs before applying over the Test-Set data sample. Figure 9 shows that the percentage score is shown in terms of Accuracy, Precision, and Recall.

**Figure 9.**
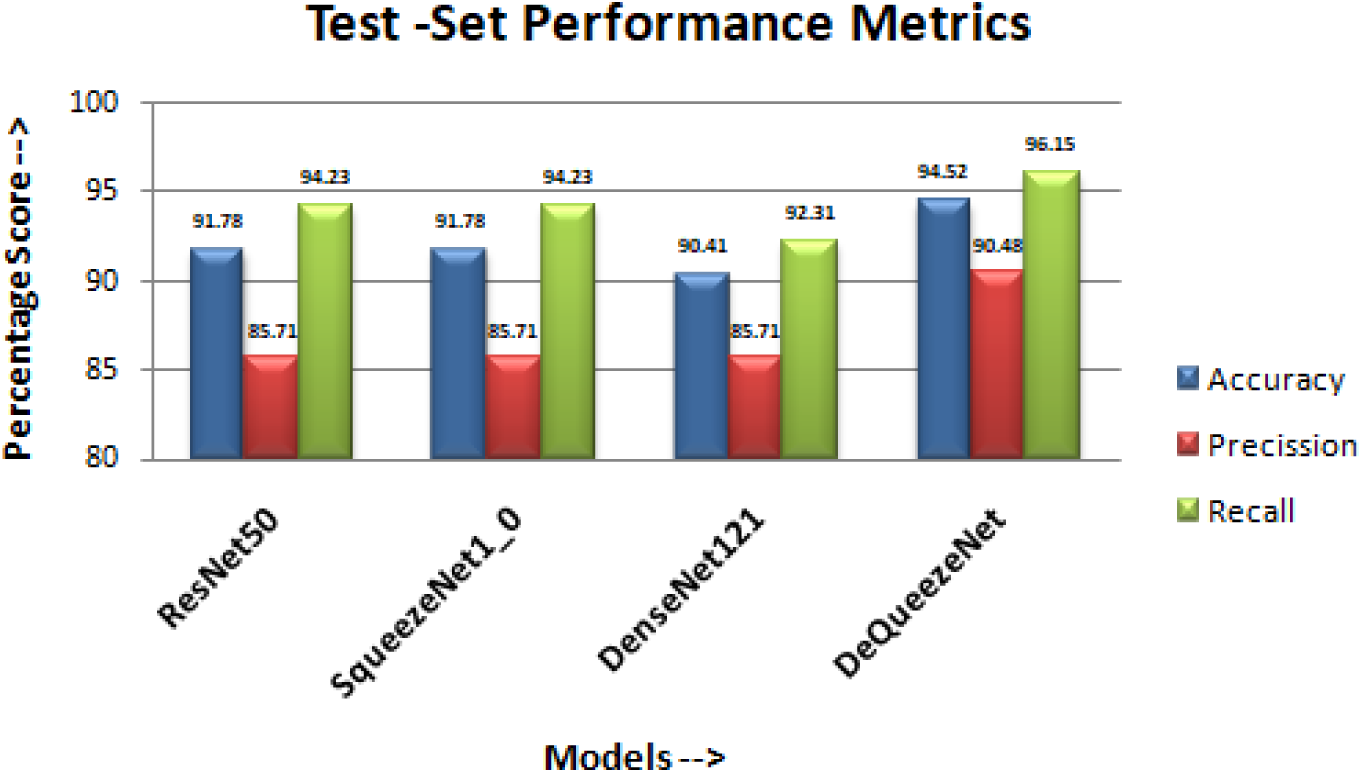
Comparative performance in terms of percentage score

Observation from figure 9, shows that the proposed model DeQueezeNet is performing better than all the other three models in terms of accuracy, precision, and recall. The performance of the DeQueezeNet is not only better than SqueezeNet1.0 and DenseNet121 but it is also better than recently proposed model ResNet50 [34] by Narin et al.

## 7. Conclusion

In this paper, we have proposed an ensemble-based DeQueezeNet model which consists of DenseNet121 and SqueezeNet1.0. The performance evaluation is done by applying the model on X-ray images to predict COVID-19 positive and negative cases.

Performance evaluation is done by considering the performance metrics which are accuracy, precision, and Recall. The confusion matrix shows that the proposed model can identify the COVID-19 positive and negative cases effectively. Suitable accuracy and high precision imply the significance of the model.

A comparative study is also done with recent work on the above-stated performance metrics. In which, it is observed that performance of the proposed model significantly better which signifies the importance of the model. And it also justifies that the proposed model is best suited for classifying the COVID-19 positive and negative cases.

## Data Availability

All data used in for the study is obtained from following two open-source datasets
i] Joseph Paul Cohen and Paul Morrison and Lan Dao
COVID-19 image data collection, arXiv:2003.11597, 2020
ii] Chest X-Ray Images (Pneumonia) by Paul Mooney on Kaggle

https://github.com/ieee8023/covid-chestxray-dataset

